# Willing everywhere but in health: a within-person national survey of artificial intelligence acceptance among 11 013 Nigerian adults

**DOI:** 10.64898/2026.09.05.26362338

**Authors:** Samson Ikenna Abanni, Agbo Uchechukwu, Muzammil Yaqub Sulaiman, Jean Omotosho, Paschaline Nchedo, Sunday Oge, Melwin Ekure, Oyari Oko Felix-Ogbonia, Uzochukwu Ofonakara, Ogodo Lucy Uzodinma

## Abstract

**Introduction:** Artificial intelligence is entering African primary care partly on the premise that populations underserved by clinicians will accept an algorithm in a clinician’s place, a premise rarely tested where it matters. We measured how far Nigerian adults separate health from every other use of AI in their lives.

**Methods:** Cross-sectional national survey of 11 013 adults across Nigeria’s 36 states and Federal Capital Territory, July–October 2025. About 60% were recruited face to face, with translation where needed, so neither literacy nor English was required to take part. Each rated willingness to use AI in nine everyday situations on a 1–5 scale, one being health advice when no doctor was available. The primary outcome was a within-person difference: health minus the respondent’s mean of the other eight. Associations were estimated by ordinary least squares with cluster-robust (CR2) standard errors by state.

**Results:** Health finished last of the nine (mean 3.10), below promoting one’s own political party (3.40). The within-person gap was −0.568 (95% CI −0.594 to −0.543; d_z_ −0.417), widening to −0.904 among the 6 924 who distinguished between domains. Keeping a clinician in view did not relieve it: 22.9% would trust AI to help doctors diagnose against 25.5% who would use it to check their own symptoms. Among barriers respondents named, the gap was wider for language (β −0.427, 95% CI −0.718 to −0.136) and unfriendly staff (−0.411, −0.616 to −0.207), narrower for missing equipment (+0.231, +0.040 to +0.423) and narrower among fluent English speakers (+0.386 per SD, +0.130 to +0.643).

**Conclusion:** Nigerian adults accept AI across their lives and withhold it from their health, most firmly where care has been unaffordable, unintelligible or unkind. No configuration tested commanded majority willingness, including AI presented as assisting a clinician; willingness belongs alongside connectivity and data in deployment planning, not assumed.

**Key questions:** *What is already known on this topic:*

- Health systems in low- and middle-income countries are being encouraged to adopt patient-facing AI, including symptom checkers positioned as a first point of contact without provider oversight, on the reasoning that people will turn to AI where access to care is poor.
- The best-identified experimental default in lay judgment is algorithm *appreciation* rather than aversion; resistance to medical AI is compensatory and can be bought off by demonstrated accuracy.
- Patient-attitude evidence is dominated by high-income settings and hospital samples; reviews and strategies that enumerate barriers to health AI in Africa do not count population willingness among them.

*What this study adds:*

- In a national sample of 11 013 Nigerian adults, health was the least acceptable of nine uses of AI, below promoting one’s own political party, with the comparison drawn within each respondent so that scale-use habits cancel.
- The discount was not explained by stakes, by inattentive answering, or by keeping a clinician in the loop, and it was not closed by digital capability.
- It was largest among respondents whose stated grievance with the health system was language or disrespect, and among the uninsured, and smallest among those whose grievance was missing equipment: the gap tracks how people were treated rather than what the clinic lacked.

*How this study might affect research, practice or policy:*

- Not one of the four framings tested, including AI presented as assisting a clinician, reached majority willingness, so willingness is a constraint to be measured before deployment rather than a barrier to be educated away.
- Willingness is cheap to measure and belongs in African health-AI strategy alongside connectivity, data and governance, and in these data the sharpest predictor of it is language.

## Introduction

Artificial intelligence is arriving in primary care with a comfortable chair already chosen for it. A 2026 *Lancet Primary Care* review records that countries are placing symptom checkers in “virtual frontdoor” roles as the first point of contact with the health system, without provider oversight, on triage accuracy the same review calls variably low to moderate. It states the operating premise plainly: in settings with restricted access to care, AI “might be increasingly seen as an alternative for some primary care needs,” because “people turn to AI when facing gaps in care access.”^1^ A companion viewpoint sets out the scenario in a near-future vignette: a mother in a remote village borrows a neighbour’s phone, describes her son’s fever in her own dialect, and an AI triage tool produces a risk assessment for later clinician review.^2^ The World Health Organization’s Foreword to its health-AI ethics guidance offers the same reasoning at the level of policy: with AI-based tools, “governments could extend health care services to underserved populations”.^3^ The African Union’s digital health strategy directs the promise at rural and remote areas in particular.^4^

The premise is clearly demand-side, but the evidence assembled behind it is almost entirely supply-side. A 2026 *Lancet Regional Health – Africa* comment by authors spanning Africa CDC and five universities enumerates what could stop AI generating public value on the continent: fragmented data ecosystems, uneven connectivity and unreliable power, limited interoperability, thin capacity to evaluate and procure, vendor lock-in, data sovereignty, cybersecurity, and training data that exclude rural and displaced groups.^5^ Whether the population will use the tools does not appear on the list. Nor does it appear among the AU’s barriers to scaling digital health, which are infrastructure, funding and workforce skills; there, trust enters once, as something privacy regulation will supply.^4^ Meanwhile deployment proceeds: a 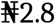 billion AI grant was distributed through Nigeria’s Federal Ministry of Communications, Innovation and Digital Economy in October 2024, and a Nigerian Healthcare AI Implementation Group was launched in September 2025.^6^ Nigerian clinicians have set out the demand-side question themselves, and predicted the answer. AI acceptance, they write, carries “social, cultural, and religious concerns,” and Nigeria has “seen apathy to other interventions in the past such as the polio vaccine, family planning, and the COVID-19 vaccine.”^7^ It has not been tested at population scale. What is known from the region comes from providers. Across six Gates Foundation AI Grand Challenges projects, 191 frontline health workers who had used generative AI tools produced 617 responses, of which 75.4% were rated enthusiastic by a panel of nine evaluators; just over half the workers were in Africa.^8^ Their open answers were less uniform than that figure suggests, with cultural and linguistic mismatch recurring among the sceptical.

Nor does the experimental literature settle it. The better-identified default in lay judgment is algorithm *appreciation*: given identical advice relabelled, people weight it more heavily when told an algorithm produced it, across tasks from estimating a stranger’s weight to forecasting romantic attraction — a result 119 judgment researchers asked to predict got backwards.^9^

Aversion is the narrower phenomenon. The label was coined for what happens after people watch an algorithm err; before seeing any error, the same participants chose the algorithm over their own judgment.^10^ Resistance in medicine specifically is real but compensatory: it is driven by a belief that one’s own case is unique, and demonstrated superior accuracy buys it off.^11 12^ Population evidence, where it exists, is high-income and mostly clinical: among 13 806 hospital patients across 43 countries, 57.6% held a generally positive attitude to health AI while 72.9% preferred physician-led decisions even at some cost in accuracy;^13^ among US adults, 65.8% had low trust in their health system to use AI responsibly, and previous experience of discrimination in care predicted lower trust still.^14^ In Australia, 9.9% of adults had used ChatGPT for health information in six months, with use concentrated among people with lower health literacy and those born outside English-speaking countries.^15^ One general-population study in the region exists. Asiedu and colleagues surveyed 672 adults across Ghana, Rwanda, Nigeria, Kenya and South Africa and found generally positive attitudes and high trust in AI for health.^16^ Their survey was conducted online in English, and to take part a respondent had to hold at least a high school diploma, attest to reading complex technical English “very well”, and already be familiar with the term AI. The authors name the restriction themselves as a limit on generalisability. That is the population most digital-health surveys reach, and it is not the population on whose behalf these deployments are argued for. What is missing is a sample recruited without those filters, and a design that asks the same person about health alongside the rest of their life, so that a reticent answer about health cannot simply be a reticent respondent.

We asked 11 013 Nigerian adults how willing they would be to use AI in nine everyday situations, one of them health advice when no doctor was available, and measured the distance each respondent placed between health and everything else. To our knowledge this is the largest general population sample on this question from a low-income or middle-income country.

## Methods

### Design and setting

Cross-sectional survey of adults resident in Nigeria, fielded from 12 July to 28 October 2025 across all 36 states and the Federal Capital Territory. Reporting follows STROBE.^17^

### Participants and recruitment

Approximately 60% of respondents were recruited in person, across every local government area of each state, by trained interviewers working from a common protocol. The in-person form was interviewer-administered: interviewers read the questions aloud and translated them for respondents who could not read or speak English, so literacy and English were not conditions of participation. The remaining respondents were recruited online through the networks of state mobilisers. All respondents were required to be resident in the state in which they were recruited and aged 18 years or older. Recruitment was quota-based by zone rather than probabilistic; the in-person arm was deliberately structured to reach people with no formal schooling and people outside salaried work, whose views are systematically absent from online-only surveys of digital technology.

### Ethics and consent

The protocol was reviewed and approved by the Directorate of Health Research and Ethical Committee, Ebonyi State Ministry of Health (EBSHREC protocol EBSHREC/112; reference EB/SMOH/HREC/6/320; committee-assigned number 232). Consent for the interviewer-administered arm was verbal: interviewers read the information sheet in the respondent’s language and recorded consent before beginning, a waiver of written consent appropriate where a substantial minority of respondents do not read. Online respondents consented before the first item. Data were de-identified before analysis.

### Participant flow

Of 11,321 submissions, 243 duplicate records identified by submission identifier were removed, and 65 records recorded as under 18 years were removed as ineligible, leaving 11,013 for analysis. All 11,013 answered all nine willingness items.

### Outcome

Each respondent rated willingness to use AI in nine everyday situations, presented as one block on a common five-point scale from 1 (very unwilling) to 5 (very willing): health advice when no doctor is immediately available; help with work or business tasks; helping a child with homework or learning; advice on important personal decisions; researching a major purchase or investment; news and current information; promoting one’s own political party; learning skills to get a job; and carrying out academic work.

The health item differs from the other eight in one respect that bears on the comparison: it is the only one that specifies the alternative, telling respondents that no doctor is immediately available. The contrast is therefore between a domain and a domain-plus-framing rather than between domains alone. We wrote it that way to put the strongest case for health AI in low-resource settings inside the question, but it means the gap cannot be attributed to the domain in isolation.

The primary outcome was the difference, within each respondent, between willingness for health and the mean of their eight other ratings. The difference has a useful arithmetic property. Any disposition that shifts a respondent’s whole row (a habit of using the top or bottom of a scale, acquiescence, a wish to appear agreeable) shifts both terms equally and cancels. What remains is the separation the respondent drew themselves.

### Other measures

Six items on self-rated digital skills were averaged into a skills score. Respondents named up to three principal barriers to healthcare in Nigeria from eight options, reported whether they held health insurance, rated their spoken and written English on five points, and answered four items placing AI at different distances from a clinician together with two items on past behaviour. A further item asked which of eight institutions they trust information from AI more than.

### Statistical analysis

We report means with 95% confidence intervals, paired contrasts between health and each other domain with Cohen’s d_z_, and proportions. Associations with the within-person gap were estimated by ordinary least squares with standard errors clustered on the 37 states and the Bell–McCaffrey (CR2) small-sample correction, with t-tests on 36 degrees of freedom; continuous predictors were standardised. The CR2 adjustment was implemented directly from the Bell–McCaffrey definition (the cluster-wise inverse square root of I − H), because the cluster-robust estimator distributed with statsmodels is CR1; the implementation is included with the analysis scripts. The model was fitted among respondents who distinguished between at least two domains, since a respondent who gave the same answer to all nine has a difference of zero by construction; because that restriction is defined on the outcome, the same model fitted on all respondents is reported alongside it, and we make no claim that does not hold in both. Analyses were conducted in Python 3 (pandas, statsmodels, scipy). No adjustment was made for multiple comparisons.

### Patient and public involvement

Patients and the public were not involved in the design, conduct or reporting of this study. Respondents were the source of the data. A plain-language summary in English and major Nigerian languages will be distributed through the recruiting networks at publication.

## Results

### Sample

Of 11,013 respondents, 49.8% were male and 47.3% female; median age was 32 years (IQR 25– 45; mean 37.4, SD 16.6). Just under half lived in urban centres (46.4%), a third in peri-urban areas (33.3%) and a fifth in rural communities (20.3%). Educational attainment spanned the range: 45.9% held a tertiary or postgraduate qualification and 18.3% had no formal schooling. A third rated their English poor or fair (33.2%). Two-thirds owned a smartphone (68.6%) and 57.2% used the internet at least daily, while one in five never used it. Only 17.9% held any health insurance. Zonal samples ranged from 870 in the South-East to 2 790 in the North-West (Table 1).

**Table 1.** Sample characteristics (N = 11 013)

|  | n (%) |
| --- | --- |
| **Age**, median (IQR) | 32 (25–45) |
| Age, mean (SD) | 37.4 (16.6) |
| *missing* | 9 |
| **Gender** |  |
| Male | 5 489 (49.8) |
| Female | 5 204 (47.3) |
| Prefer not to say | 320 (2.9) |
| **Residence** |  |
| Urban | 5 114 (46.4) |
| Peri-urban | 3 666 (33.3) |
| Rural | 2 233 (20.3) |
| **Highest education completed** |  |
| Tertiary | 4 272 (38.8) |
| Secondary | 2 900 (26.3) |
| No formal education | 2 014 (18.3) |
| Primary | 1 049 (9.5) |
| Postgraduate | 778 (7.1) |
| **Self-rated English** |  |
| Excellent | 1 046 (9.5) |
| Very good | 2 622 (23.8) |
| Good | 3 688 (33.5) |
| Fair | 1 893 (17.2) |
| Poor | 1 764 (16.0) |
| **Health insurance** |  |
| Yes | 1 969 (17.9) |
| No | 7 659 (69.5) |
| Don't know | 1 385 (12.6) |
| **Internet use** |  |
| Multiple times a day | 2 830 (25.7) |
| Daily | 3 468 (31.5) |
| A few times a week | 1 140 (10.4) |
| A few times a month | 770 (7.0) |
| Rarely | 601 (5.5) |
| Never | 2 204 (20.0) |
| **Owns a smartphone** | 7 558 (68.6) |
| **Geopolitical zone** |  |
| North-West | 2 790 (25.3) |
| South-West | 2 436 (22.1) |
| South-South | 1 649 (15.0) |
| North-East | 1 628 (14.8) |
| North-Central | 1 613 (14.6) |
| South-East | 870 (7.9) |
| *zone not recorded* | 27 (0.2) |

### Willingness across nine domains

Six of the nine domains fell within 0.11 points of one another: academic work (3.81), news and current information (3.76), helping a child with homework (3.75), learning skills for a job (3.74), work or business tasks (3.72), and researching a major purchase or investment (3.70). Two sat below that band: advice on important personal decisions (3.50) and promoting one’s own political party (3.40). Health was last, at 3.10, and it was the only item that specified what the alternative would be: respondents were asked how willing they would be to use AI for health advice *when no doctor was immediately available* (Figure 1, Table 2).

**Table 2.**
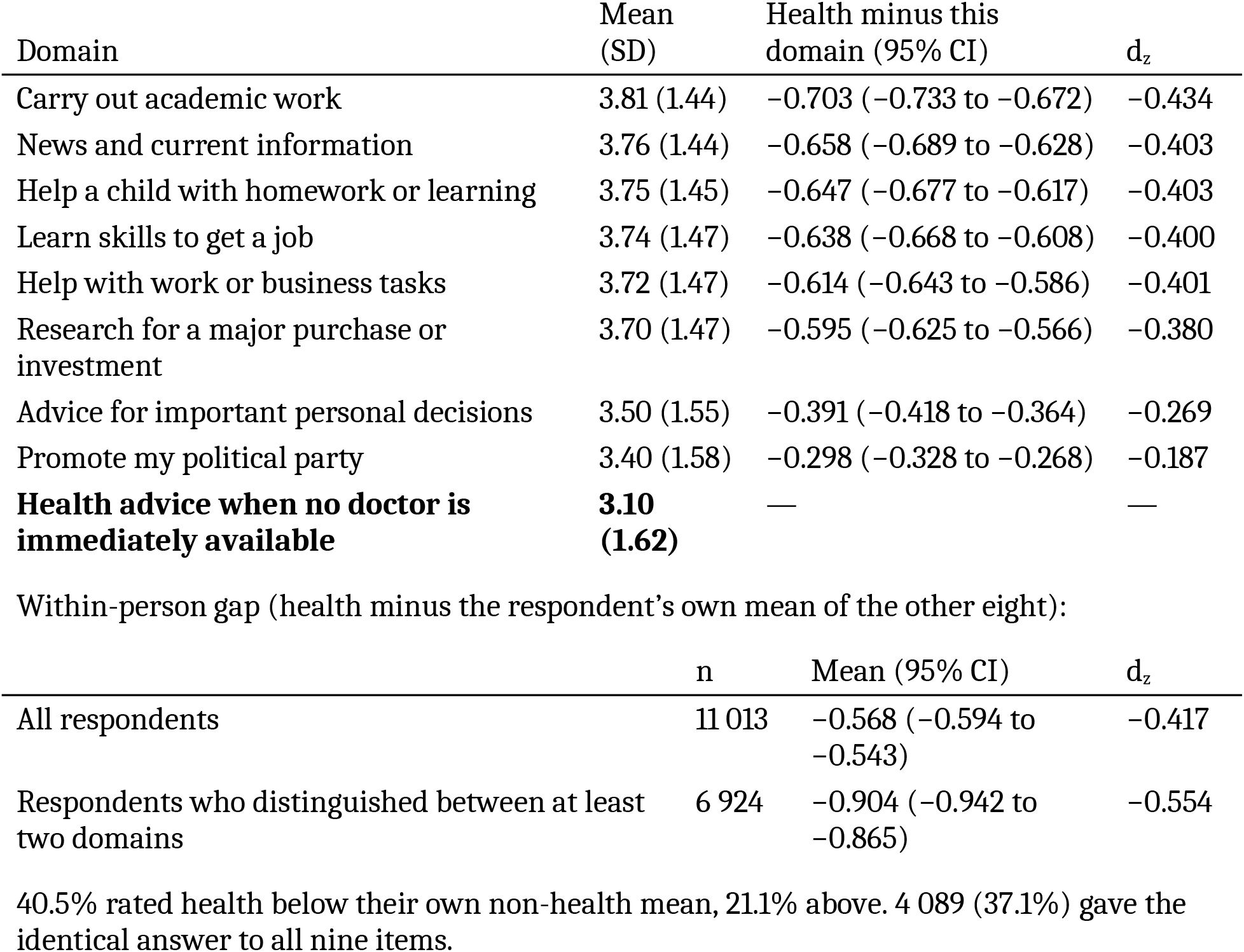
Willingness to use AI by domain, and paired contrast with health (N = 11 013) Rated 1 (very unwilling) to 5 (very willing).

| Domain | Mean (SD) | Health minus this domain (95% CI) | d<sub>z</sub> |
| --- | --- | --- | --- |
| Carry out academic work | 3.81 (1.44) | -0.703 (-0.733 to -0.672) | -0.434 |
| News and current information | 3.76 (1.44) | -0.658 (-0.689 to -0.628) | -0.403 |
| Help a child with homework or learning | 3.75 (1.45) | -0.647 (-0.677 to -0.617) | -0.403 |
| Learn skills to get a job | 3.74 (1.47) | -0.638 (-0.668 to -0.608) | -0.400 |
| Help with work or business tasks | 3.72 (1.47) | -0.614 (-0.643 to -0.586) | -0.401 |
| Research for a major purchase or investment | 3.70 (1.47) | -0.595 (-0.625 to -0.566) | -0.380 |
| Advice for important personal decisions | 3.50 (1.55) | -0.391 (-0.418 to -0.364) | -0.269 |
| Promote my political party | 3.40 (1.58) | -0.298 (-0.328 to -0.268) | -0.187 |
| **Health advice when no doctor is immediately available** | **3.10 (1.62)** | — | — |

Within-person gap (health minus the respondent's own mean of the other eight):
|  | n | Mean (95% CI) | d<sub>z</sub> |
| --- | --- | --- | --- |
| All respondents | 11 013 | -0.568 (-0.594 to -0.543) | -0.417 |
| Respondents who distinguished between at least two domains | 6 924 | -0.904 (-0.942 to -0.865) | -0.554 |

40.5% rated health below their own non-health mean, 21.1% above. 4 089 (37.1%) gave the identical answer to all nine items.

**Figure 1.**
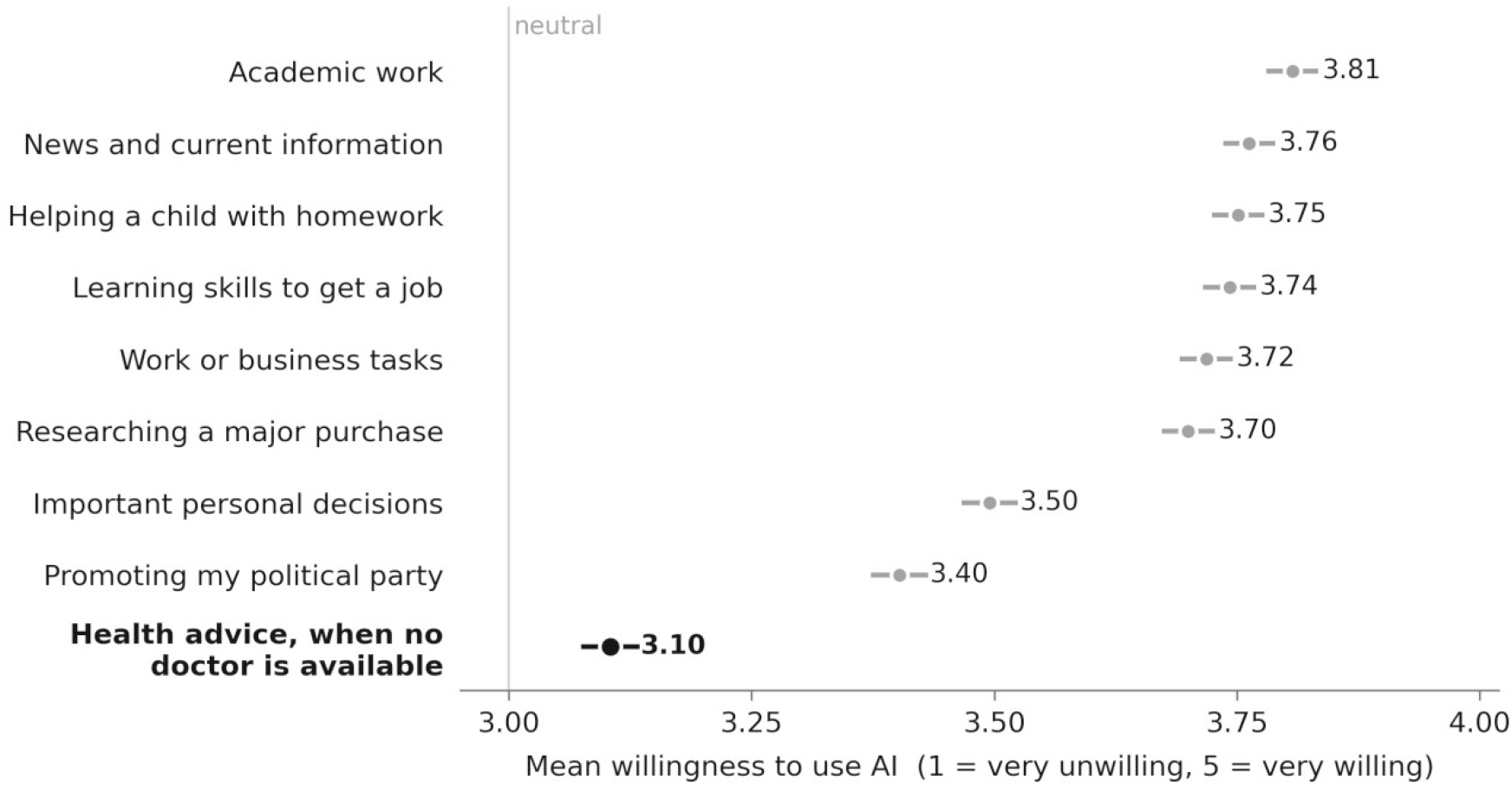
Willingness to use artificial intelligence, by domain (N = 11 013). Mean willingness across nine everyday situations, rated 1 (very unwilling) to 5 (very willing). Horizontal bars are 95% confidence intervals; at this sample size they are narrower than the markers. Health, in bold, is the only item that specified the alternative — advice sought when no doctor is immediately available — and the only one within a tenth of a point of the neutral midpoint.

### The within-person gap

Because every respondent rated all nine domains, the comparison can be drawn within the person. On that measure the mean gap was −0.568 (95% CI −0.594 to −0.543; d_z_ −0.417). Of all 11 013 respondents, 40.5% rated health below their own non-health average and 21.1% above.

### It is not about stakes

The obvious reading is that health is simply higher-stakes, and that people withhold AI where error is costly. Two of the eight comparison domains are themselves consequential, which allows the reading to be tested. Willingness for health fell 0.391 points below willingness for advice on important personal decisions (95% CI −0.418 to −0.364; d_z_ −0.269) and 0.595 points below researching a major purchase or investment (−0.625 to −0.566; d_z_ −0.380). It also fell 0.298 points below willingness to use AI to promote one’s own political party (−0.328 to −0.268). Health was rated below every one of the eight domains, in every case at p<0.001. Respondents were more willing to let an algorithm shape their politics than their treatment.

### It is not an artefact of undifferentiated answering

A second reading is measurement rather than attitude. It runs the wrong way. 4 089 respondents (37.1%) gave the identical answer to all nine items; for them the difference is zero by construction, which pulls the sample mean toward zero. Among the 6 924 who distinguished between at least two domains the gap was −0.904 (95% CI −0.942 to −0.865; d_z_ −0.554).

Undifferentiated answering conceals this finding rather than producing it, and −0.568 is a lower bound.

### Keeping a clinician in view does not relieve it

Four items placed AI at different distances from a clinician, and all 11 013 respondents answered them. 2 520 respondents (22.9%) would trust AI to help doctors make better diagnoses, slightly fewer than the 2 806 (25.5%) who would use AI to check their own symptoms before deciding whether to see a doctor at all. 23.9% would hand an AI tool their laboratory results to interpret, and 21.2% would trust AI-generated advice in public health matters such as a vaccination campaign or a disease outbreak. The four framings, spanning assistance to substitution, ran within 4.3 percentage points of one another. Reported behaviour sat lower: 14.7% had at some point asked an AI tool for health advice instead of seeing a provider, and 18.2% had shared sensitive health information with one: medical history, HIV status, mental health details.

### Where people go, and whom they would replace

Asked where they turn first when they need health information, 36.5% named a doctor or nurse, 19.2% family or friends and 16.4% a pharmacy. Google drew 9.0%. AI tools drew 6.5% (715 respondents), behind traditional healers at 8.4% (924). Presented with a scenario of persistent headache and mild fever, 24.5% said they would follow ChatGPT’s recommendation; asked about buying medicine for a child’s persistent cough on AI advice, 20.9% said it would influence them.

This unwillingness is specific to medicine rather than a general preference for institutions over machines. Asked which sources they trust information from AI more than, 58.4% named the Nigerian government. Only 22.3% named the Nigerian healthcare system, 26.3% the judiciary, 22.7% the banks, 21.6% international technology companies and 20.3% the news media (n = 10 018 answering). Trust in government was itself the lowest of the institutions measured (mean 2.85 of 5) and trust in the healthcare system among the highest (3.53). Nigerians in this sample would take an algorithm over their government. They would not take one over their doctor.

### What the gap tracks

The gap tracked how respondents described the failures of the health system, and not in the direction scarcity would predict. In a model adjusted for age, sex, education, English fluency, digital skills and the eight barrier indicators, with cluster-robust standard errors by state, naming language barriers as a principal barrier to care was associated with a gap 0.427 points wider (95% CI −0.718 to −0.136) and naming unfriendly staff 0.411 points wider (−0.616 to −0.207); respondents without health insurance carried a gap 0.276 points wider (−0.532 to −0.021). Naming a lack of equipment ran the other way and the gap was narrower (+0.231, +0.040 to +0.423); a lack of specialists, long distances, long waiting times and poor quality of care were not associated with the gap in either direction (Table 3). Each of these associations held in the same model fitted on all respondents.

**Table 3.**
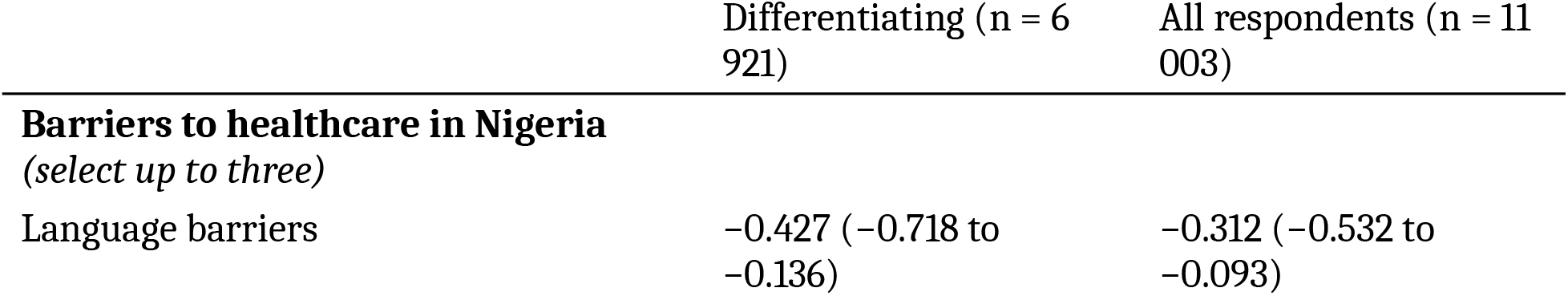
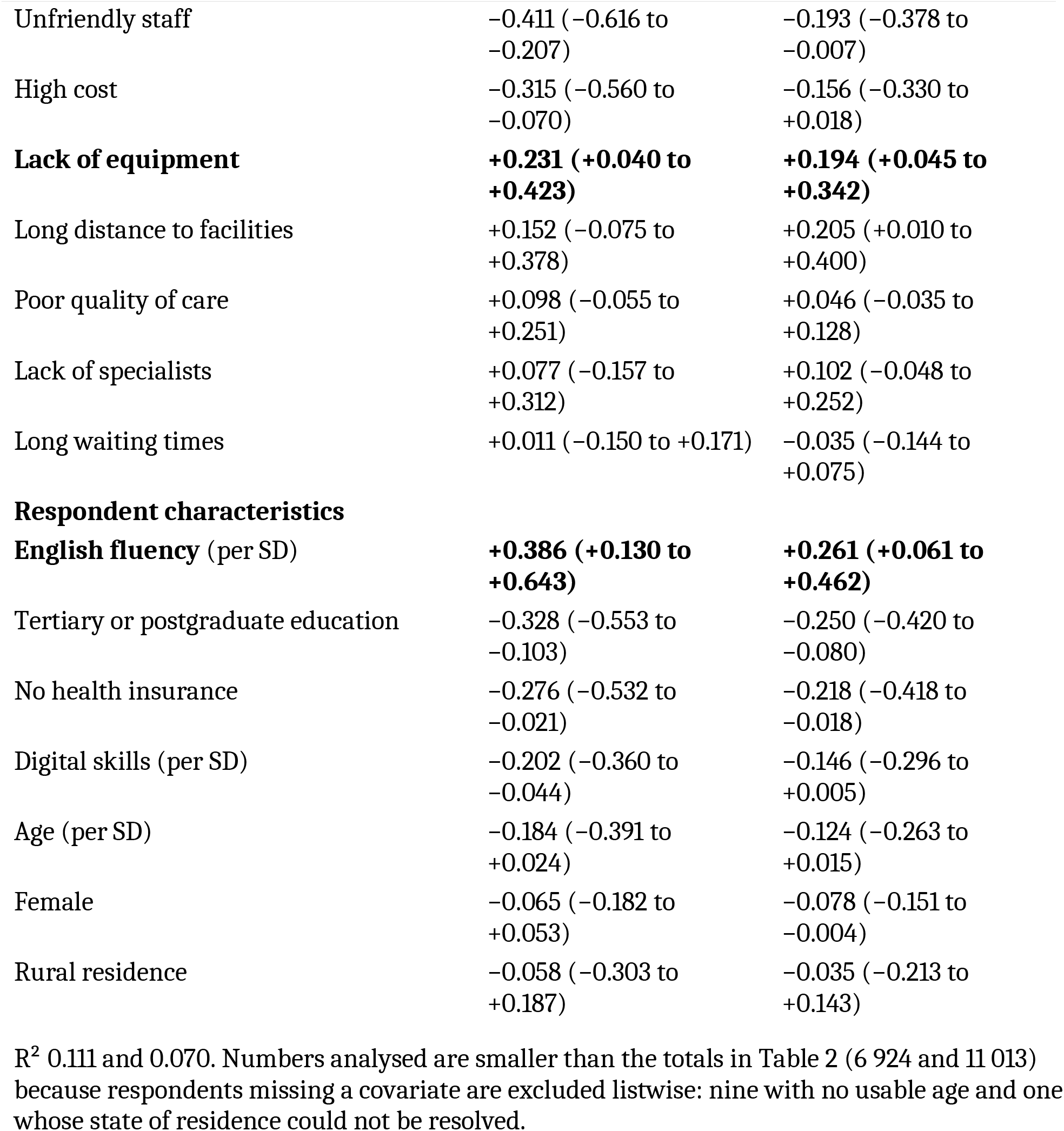
Adjusted associations with the within-person health gap. Ordinary least squares. Standard errors cluster-robust by state with the Bell–McCaffrey (CR2) correction, t-tests on 36 degrees of freedom, 37 state clusters. Continuous predictors standardised. Negative coefficients indicate a **wider** health gap. The main column is fitted among respondents who distinguished between domains; because that restriction is defined on the outcome, the same model fitted on all respondents is shown alongside.

### The gap follows language

The gap was widest among respondents least able to conduct their care in English. It ran −0.76 among those who rated their English poor, −0.77 fair and −0.68 good, and roughly a third of that, −0.26 and −0.26, among those who rated it very good or excellent. Undifferentiated answering was roughly constant across levels of fluency (40%, 36%, 33%, 40% and 40% from poor to excellent), so the gradient is not an artefact of who was willing to distinguish between domains. In the adjusted model English fluency was the strongest protective term (+0.386 per SD, 95% CI +0.130 to +0.643), against language barriers as the strongest widening one. What predicted the health discount was whether a respondent could expect to be understood inside the clinic.

## Discussion

Nigerian adults are not averse to artificial intelligence; they are averse to it in the one domain where it is most often offered to them in place of a person. Across nine everyday uses rated by the same 11,013 people, health finished last: below advice on important personal decisions, below research for a major purchase, and below promoting one’s own political party — and the health item was the only one that told respondents no doctor was available. The discount is not manufactured by inattentive answering: respondents who gave every item the same score have a difference of zero by construction, and among those who distinguished between domains the gap widens from −0.57 to −0.90. It is not relieved by keeping a physician in view. And it falls heaviest on the people whose experience of care has been worst. We call the pattern the health exception.

The experimental literature would not have predicted this, though the comparison needs care. Those studies measure how much weight people give to identical advice labelled algorithmic or human, in judgment tasks. We measure stated willingness to adopt AI across domains of life. What our data run against is the expectation imported from that literature, not its findings. The best-identified estimate of how lay people treat algorithmic advice is appreciation, not aversion.^9^ The famous aversion finding is narrower than its reputation. It was named for the effect of seeing an algorithm err, and in those same experiments participants who had seen no error chose the algorithm over their own judgment, a distinction the paper is routinely cited past.^10^ Resistance to medical AI in particular is compensatory: demonstrated superior accuracy buys it off, a caveat none of the twenty-five-plus papers then citing it had noted.^11 12^ Our respondents met none of the conditions under which aversion is predicted. They saw no algorithm err, were not weighing a machine against their own judgment, and were not domain experts protecting a professional prerogative. They discounted AI anyway, and only for health.

The framing on which most deployment guidance rests did not survive contact with these data. Resistance to medical AI has been shown to disappear when the automated provider merely provides input to a physician’s decision rather than replacing one, a result obtained by manipulating that framing within a single item.^11^ Our two nearest items ran the other way.

Willingness to trust AI to help doctors make better diagnoses (22.9%) sat below willingness to use AI to check one’s own symptoms before deciding whether to see a doctor (25.5%): 463 respondents endorsed the assistance framing but not self-triage, 749 the reverse, a paired difference of 2.60 percentage points (95% CI 1.98 to 3.22; McNemar χ^2^ = 67.0, p<0.001). Two differently-worded items are not a controlled framing manipulation, so this is not a failed replication; it is a demonstration that in this population the assistance framing carried no premium, and if anything a small penalty. That marks the boundary of the framework rather than refuting it, because the manipulation works by answering a doubt about the algorithm, and a reservation that is not about the algorithm gives it nothing to answer.

The distinction that matters for policy is between relative and absolute acceptance. A preregistered conjoint study of 3 000 US adults found clinician presence raised the probability of choosing a visit by 18.4 percentage points (95% CI 17.3 to 19.5).^18^ That design asks people to choose between AI-involving scenarios and measures preference among configurations. Ours asks about each configuration on its own. A person can prefer clinician-supervised AI to unsupervised AI while being unwilling to use either, and where the question is whether a population will take up a service at all, it is the absolute quantity that binds.

Respondents naming a lack of specialists, long distances or long waiting times carried no larger gap than anyone else, and those naming a lack of equipment carried a smaller one. Those naming language or unfriendly staff carried larger gaps, as did respondents without health insurance; naming cost was associated with a larger gap among respondents who distinguished between domains, but that association did not survive the full-sample model. The pattern runs against scarcity and towards treatment, and we can describe the health exception with more confidence than we can explain it.

One reading is the competence–warmth division the trust literature has begun to draw. In a randomised field experiment among 15 000 users of a mobile health app, disclosing AI reduced uptake of suggested behaviour change relative to the same advice from a human expert; the deficit lay in affective rather than cognitive trust, and it recovered when the tool was paired with human expertise, made transparent, or framed to convey warmth.^19^ On that reading a machine may stand in for a missing ultrasound but not for a person who listens, and two observations elsewhere are consistent with it: among US adults, prior experience of discrimination in care predicted lower trust that the health system would use AI responsibly,^14^ and language, the sharpest form of the pattern here, appears again from the provider side, where a health worker in that study described AI translations as “often too literal” and as missing “the nuances of our language.”^8^ Two other readings fit these data as well. The barrier items ask what respondents consider the biggest barriers to healthcare in Nigeria rather than what they have personally met, so naming language or unfriendly staff may index a respondent’s position in society rather than an encounter. And a general disposition to distrust institutions could produce both a harsher account of the health system and a lower willingness to take health advice from a machine, with no causal path running between them. A cross-sectional survey cannot separate the three, and the associations in Table 3 should be read as a description of who carries the gap rather than an account of why.

Traditional healers outranked AI tools as a first source of health information, 924 respondents to 715. Whatever the clinical merits of that ordering, it is the ordering the population reports, and it runs against the premise that populations underserved by clinicians will leapfrog to algorithms. A related assumption also fails: that people will tell a machine what they will not tell a person. In a between-subjects experiment comparing face-to-face, human-mediated and chatbot consultations, the chatbot drew the least willingness of the three to disclose sensitive health information, trust concerns overriding any disinhibition effect.^20^ In our sample 18.2% had ever shared sensitive health information with an AI tool.

Placed beside high-income evidence, the Nigerian figure is not exotic. Australian adults who had used ChatGPT for health information rated their trust in it at 3.1 of 5;^15^ our national mean willingness for health AI is 3.10 of 5. The constructs differ: theirs is trust among users, ours willingness across the whole adult population. The coincidence is still enough to unsettle any assumption that the health exception is a low-income problem to be educated away. What does differ is direction. In Australia, health-AI use runs towards the underserved: lower health literacy, migrants, non-English speakers.^15^ Among US adults, trust in AI diagnostic capability was higher among the uninsured than the insured.^21^ Ours runs the other way, and the counterfactual explains why: in a high-income system, being uninsured means exclusion from care that exists and works, so AI substitutes for an unaffordable option. In Nigeria 69.5% of our sample reported having no insurance; the alternative to a clinician is frequently not an expensive clinician but none at all, and a tool that cannot be understood or trusted does not fill that space. The *Lancet* review that names the inverse care law as a risk of AI implementation frames it as a supply-side problem: good tools least available where need is greatest.^1^ Our data give it a demand-side form: those in greatest need are also least willing.

Three features carry the primary finding. The comparison is drawn within the person, so any disposition that shifts a whole row cancels in the difference. That also disarms the strongest methodological objection to stated-preference work in this literature, that hypothetical elicitation inflates apparent algorithm aversion relative to real judgments.^10^ The health item stipulated that no doctor was available. If the absence of a clinician were what unlocks acceptance, this is the item on which willingness should have been highest; it was the lowest of the nine. And the in-person, interviewer-administered arm reached 2 014 respondents with no formal schooling and 3 657 who rate their English poor or fair, whose views are structurally absent from online surveys about digital technology.

The limitations are real and we name them with their direction. The design is cross-sectional; we cannot say whether willingness moves with exposure, so these figures describe one moment in a fast-moving technology and should be re-measured rather than extrapolated forward.

Stated intention is not behaviour, though two items measured behaviour directly and pointed the same way. The within-person difference cancels hypothetical-elicitation bias only if that bias is domain-invariant; if people overstate willingness more readily for shopping than for health, the gap is confounded, and these data cannot rule that out. Quota sampling is not probability sampling and the zonal quotas were unequal, which unsettles any national point estimate; we report none, and the primary outcome is a comparison each respondent makes with themselves. The sample is also far more educated than the Nigerian adult population: 45.9% held a tertiary or postgraduate qualification, against 13.7% of women and 20.5% of men aged 15–49 with more than secondary education in the 2024 Nigeria Demographic and Health Survey.^22^ We cannot sign the resulting bias. Unadjusted, respondents with no formal schooling carried the widest gap of any education group; in the adjusted model, tertiary education was associated with a wider one. The barrier items record perceptions of the health system rather than reports of personal experience, which weakens any causal reading of the associations in Table 3 in either direction. Just over a third of respondents (37.1%) gave the same answer to all nine willingness items, which fixes their difference at zero and attenuates the overall gap; the adjusted model was therefore fitted among respondents who distinguished between domains, with the full sample reported alongside, and we make no claim that does not survive both. The analyses were not pre-registered; the code that produced every number reported here is deposited, so the specification can be read against the results.

The implication we draw is narrow. None of the configurations we asked about commanded majority willingness, and the one that placed AI alongside a clinician did no better than the one that placed it in front of the patient, so this study endorses no patient-facing deployment. What it does separate is degree. Respondents whose complaint about the health system was a missing machine discounted AI least; those whose complaint was being unheard or badly treated discounted it most, and the second group is the one these tools are most often justified as serving. That ordering suggests AI directed at what a facility materially lacks will meet less resistance than AI offered in place of the encounter. It is a claim about relative resistance, not about acceptance. Willingness is measurable and cheap to measure, and it belongs in African health-AI strategy alongside data, power and procurement. Language coverage is the first place an implementer could act on it.

## Declarations

### Contributors

SIA conceived the study, led the analysis and drafted the manuscript. MYS, AU and JO contributed to study design, data-collection oversight and interpretation. PN supported community recruitment and field implementation. SO contributed to clinical interpretation and revised the manuscript critically for important intellectual content. ME, OLU, OOF-O and UO contributed to data collection oversight and revised the manuscript critically. All authors had full access to the analytic data, take responsibility for the integrity of the data and the accuracy of the analyses, agree to be accountable for all aspects of the work, and approved the final manuscript.

## Acknowledgements

We thank the field teams across the 36 states and the Federal Capital Territory who collected the survey, and the 11 013 Nigerian adults who gave their time.

## Funding

This work received no external funding.

## Competing interests

None declared.

## Ethics approval

Directorate of Health Research and Ethical Committee, Ebonyi State Ministry of Health — EBSHREC protocol EBSHREC/112, reference EB/SMOH/HREC/6/320, committee-assigned number 232. Verbal informed consent was obtained from all participants.

## Patient and public involvement

Patients and the public were not involved in the design, conduct or reporting of this research.

## Data availability

The de-identified analytic dataset, its codebook, and the script that derives it from the raw survey export are openly available in Zenodo at **https://doi.org/10.5281/zenodo.22313218**. Identifiable raw survey records are restricted under the ethics approval (EBSHREC/112) and are not shared.

## Use of artificial intelligence

Anthropic Opus 5 was used in language editing and analysis scripting. It was not used to design the study, to collect data, or to decide which analyses to report, and it is not an author. The authors take full responsibility for the whole of the content, including the parts drafted with AI assistance.

## Notes

### Competing Interest Statement

The authors have declared no competing interest.

### Author Declarations

The protocol was reviewed and approved by the Directorate of Health Research and Ethical Committee, Ebonyi State Ministry of Health (EBSHREC protocol EBSHREC/112; reference EB/SMOH/HREC/6/320; committee-assigned number 232)

